# The Next Evolution of Hereditary Cancer Genetic Testing Service Delivery: A Descriptive Study of a Primary Collaborative Care Model for Telegenetics

**DOI:** 10.1101/2025.10.21.25338486

**Authors:** Daniel Chavez-Yenter, Kevin Oeffinger, Brian Egleston, Elisabeth McCarty Wood, Sarah Howe, Sarah Brown, Janice Christiansen, Cara Cacioppo, Michelle Weinberg, Elena Elkin, Linda Fleisher, Rajia Mim, Santina Hernandez, Demetrios Ofidis, Dominique Fetzer, Tara O. Henderson, Angela Bradbury

**Author notes:** **<u>Corresponding author:</u>** Daniel Chavez-Yenter, PhD, Department of Medicine, Division of Hematology/Oncology, Department of Medical Ethics & Health Policy, 3400 Civic Center Blvd., Philadelphia, PA 19104. Senior Authors.

## Abstract

Clinical genetic testing demand has increased in the era of precision medicine. However, availability of cancer genetic services remains limited in the US, prompting a rise in telehealth delivery. This report describes the Penn Telegenetics Program experience using a local healthcare provider collaborative model. From 2018-2025, 473 providers (89.4%) successfully registered. Providers were predominately MD/DO licensed (85.0%). Family medicine was the most frequent speciality (54.3%), followed by internal medicine (20.9%) and clinical oncology (11.8%). Most providers were in suburban locations by zip codes (56.2%), 20.1% were in rural zip codes. Only 56 providers declined collaboration (10.6%); the most common reasons reported were preferring local genetic services (22.5%) and not being comfortable as the ordering provider (22.5%). Our data demonstrate that most local providers are willing to collaborate with a centralized telegenetics program. Refining procedures to increase collaborative care and engagement may provide opportunities to increase access to cancer genetic testing.

---

Given the exponential growth of genetic testing services and the limited availability of genetic counselors (GCs) in many areas, there has been an increased interest in providing clinical genetic services via telehealth.^1–3^ Many genetic programs offer remote services within their state, but broader provision of services are limited by state licensure requirements and limited reimbursement for genetic counseling and telehealth services. Additionally, successful implementation of genetic testing includes changes to screening and risk reduction based on test results, which ultimately requires collaboration with local primary care providers (PCPs)^4^.

The Penn Telegenetics Program was founded in 2012 to expand genetic services to community practices without access to genetic counselors.^5^ Initially, remote services were provided on-site in collaboration with specific practices and providers.^6^ As the program expanded, we sought to provide services nationally to patients who meet clinical guidelines for genetic testing and have limited access to genetic services. Further, we sought to allow patients to self-refer to the program. To achieve this expanded access, we developed the in-home collaborative PCP or local provider (PCP/LP).^7^ This delivery model addresses licensure requirements (the GC and physician must be licensed in the state that the patient resides), provides care in the home and allows patients across the U.S. - regardless of location or proximity to a participating clinic - to access remote telegenetic services with their PCP/LP.^7^

This in-home collaborative PCP/LP model of care has been used across multiple research studies providing remote cancer genetic services across the US through the Penn Telegenetics Program (see **Table 1**)^7–9^. In this brief report, we report procedures and our initial experience implementing the in-home collaborative PCP/LP model across four research studies.

**Table 1.** Characteristics of providers who agree to registration and collaboration with the Penn Telegenetics Program.

| **Provider Characteristics** | **ENGAGE**<br>**(n=127)**<br>**CT.Gov #**<br>**NCT04455698** | **Patient Access**<br>**Program (n=107)** | **eREACH1**<br>**(n=24)**<br>**CT.Gov #**<br>**NCT04353973** | **eREACH2**<br>**(n=215)**<br>**CT.Gov #**<br>**NCT04353973** | **Total**<br>**(n=473)** |
| --- | --- | --- | --- | --- | --- |
| *Gender<sup>NS</sup>* | n, % | n, % | n, % | n, % | n, % |
| Male | 62 (48.8) | 46 (43.0) | 10 (41.7) | 89 (41.4) | 207 (43.8) |
| Female | 65 (51.2) | 61 (57.0) | 14 (58.3) | 126 (58.6) | 266 (56.2) |
| *Type of Provider<sup>NS</sup>* |  |  |  |  |  |
| MD/DO | 102 (80.3) | 92 (86.0) | 24 (100) | 184 (85.6) | 402 (85.0) |
| NP | 17 (13.4) | 11 (10.3) | 0 (0) | 19 (8.8) | 47 (9.9) |
| PA | 8 (6.3) | 4 (3.7) | 0 (0) | 12 (5.6) | 24 (5.1) |
| *Specialty\*\** |  |  |  |  |  |
| Family Medicine | 81 (63.8) | 65 (60.7) | 6 (25.0) | 105 (48.8) | 257 (54.3) |
| Internal Medicine | 38 (29.9) | 15 (14.0) | 3 (13.0) | 43 (20.0) | 99 (20.9) |
| Clinical Oncology<br>(Medical,<br>Surgical,<br>Radiation) | 4 (3.1) | 8 (7.5) | 15 (62.5) | 29 (13.5) | 56 (11.8) |
| OB/GYN | 3 (2.4) | 10 (9.3) | 0 | 34 (15.8) | 47 (9.9) |
| Medicine<br>Subspecialty<br>(Cardiology,<br>Gastroenterology,<br>Nephrology,<br>Pulmonology) | 3 (2.4) | 3 (2.8) | 0 | 8 (3.7) | 14 (3.0) |
| Other Speciality<br>(Urology,<br>Dermatology,<br>Surgery, etc) | 1 (.78) | 2 (1.9) | 0 | 5 (2.3) | 8 (1.7) |
| Unknown | 0 | 1 (.93) | 0 | 0 | 1 (.21) |
| *Type of Practice\*\** |  |  |  |  |  |
| Groups Practice<br>(3 or more<br>providers) | 78 (61.4) | 45 (42.1) | 16 (47.1) | 101 (47.0) | 240 (51.0) |
| Solo or two-<br>person practice | 26 (20.5) | 16 (15.0) | 0 (0) | 24 (11.2) | 66 (14.0) |
| Multi-speciality<br>Group Practice | 10 (7.9) | 33 (30.8) | 4 (11.8) | 61 (28.4) | 108 (22.8) |
| Academic<br>Faculty Practice | 3 (2.4) | 4 (3.7) | 2 (5.9) | 18 (8.4) | 27 (5.7) |
| Federally<br>Qualified Health<br>Center | 4 (3.1) | 0 (0) | 0 (0) | 5 (2.3) | 9 (1.9) |
| Other | 0 (0) | 2 (1.9) | 0 (0) | 1 (.46) | 3 (.63) |
| Unspecified | 6 (4.7) | 7 (6.5) | 2 (5.9) | 5 (2.3) | 20 (4.2) |
| *Practice Setting*<br>*Location\*\* ^* |  |  |  |  |  |
| Suburban | 76 (59.8) | 51 (47.7) | 14 (58.2) | 125 (58.1) | 266 (56.2) |
| Rural | 18 (14.2) | 37 (34.6) | 6 (16.7) | 34 (15.8) | 95 (20.1) |
| Urban | 24 (18.9) | 14 (13.1) | 3 (12.5) | 49 (23.0) | 90 (19.0) |
| Unspecified | 9 (7.1) | 5 (4.7) | 1 (4.2) | 8 (3.7) | 23 (4.9) |
| *Region of the US\*\*, ^* |  |  |  |  |  |
| Northeast | 29 (22.8%) | 62 (57.9%) | 16 (66.7%) | 143 (66.5%) | 250 (52.9%) |
| Midwest | 27 (21.3%) | 16 (15.0%) | 3 (12.5%) | 15 (7.0%) | 61 (12.9%) |
| Southeast | 29 (22.8%) | 17 (15.9%) | 1 (4.2%) | 28 (13.0%) | 75 (15.9%) |
| Southwest | 20 (15.7%) | 6 (5.6%) | 1 (4.2%) | 7 (3.3%) | 34 (7.2%) |
| West | 21 (16.5%) | 6 (5.6%) | 3 (12.5%) | 22 (10.2%) | 52 (11.0%) |
| *Provider Refusals^^* | 12 (8.6%) | 5 (4.5%) | 0 (0%) | 35 (14.0%) | 56/529 (10.6%) |

NS=non-significant; \*Respondents could have selected more than one, and percentage calculated by Study Program; \*\*p<.001
^self-reported by Zipcode (US Census); ^^not included in the table totals; percentage of study not total refusals

Our procedures have been informed by the ENGAGE^7^ Study PCP Advisory Board, but applied across all the research studies. Patients enrolling on each of the four research studies are requested to provide the name and contact information for their local PCP/LP. The research team contacts the office to confirm the mutual patient and then faxes a registration packet, which includes an informational flyer and the provider registration form (which can be completed by office staff). The one-page flyer describes the Telegenetics program, what we are asking of local providers and how they can contact the team for any questions. The registration form requests: the providers name, provider type (MD, DO, NP, PA, etc), provider speciality, practice contact information and address, gender, and practice type. Registration is considered part of clinical care and not direct engagement in research, as these procedures could be utilized if we were providing clinical billable services.

If the registration form is not returned, the research team contacts the providers office to confirm receipt and remind the office that testing can’t proceed without the provider registration. If provider’s decline, the Penn telegenetics physician attempts to contact the local provider directly to discuss the value of testing and address any concerns. Reasons for declining registration, as reported by the office staff or provider, are recorded. If a provider declines or can’t be reached, patients are contacted to see if they would like to provide a second local provider.

Patients complete pre-test counseling according to individual study procedures. The GC coordinates with the patient to select the appropriate testing and to order the test, including the local provider on the test requisition with the GC. Genetic test results are shared with the patient by the Penn GC via telehealth or digital platforms according to study protocols. All patients have the option to speak directly with a GC, even if randomized to a digital visit. GCs are licenced in US states as required by state licensure laws. The GC pre-test counseling visit note, disclosure visit note and test results are faxed to the local provider.

Registration attempts and successful registrations are recorded in study databases. Categorical data includes provider type, speciality, practice location, gender, and practice type. Reasons for declining registration and testing are recorded when available. Categorical data are presented descriptively. Chi-square tests were used to test across categorical data and Generalized Estimating Equation models for repeated measures by study in Table 1. Qualitative reasons for refusal were open coded by two investigators (DCY, AB) for analysis.

Across the studies employing the in-home collaborative PCP/LP model (2018-2025), 529 providers were contacted, with 473 providers (89.4%) successfully registered (**Table 1**). Providers were predominately MD/DO licensed (85.0%). Most came from a group practices (51.0%) or multi-specialty practices (22.8%). Family medicine was the most frequent speciality (54.3%), followed by internal medicine (20.9%) and oncology (11.8%). Most providers were in suburban locations by zip codes (56.2%) and 20.1% were in rural areas. Only 4 states, Nebraska, Oregon, South Dakota, and Hawaii did not have any registered providers. Noted significant differences across studies in provider characteristics were in practice setting location (p=.005), provider speciality (p<.001), type of practice (p<.001), and region setting (p<.001). Provider specialities varied by study, reflecting the populations recruited. For example, ENGAGE had higher representation among PCPs, while eREACH1 (recruiting patients with metastatic cancer) included higher representation from oncology.

Although most local providers were willing to register and collaborate with the program to facilitate genetic testing for their patients, 10.6% of providers declined registration resulting in their patient not receiving genetic testing through our program (**Table 2**). The most frequent reasons for declining registration were the provider’s preference for local genetic testing services (22.5%) and not being comfortable being on the genetic testing requestion form (22.5%). Some providers reported they needed to see or speak with the patient before testing (14.1%). In some cases, patients reported that they had spoken with their local provider and decided not to pursue genetic testing (14.1%). Other reasons for failed registrations included the provider no longer being at the practice or not being able to reach the provider after multiple attempts.

**Table 2.** Reasons for providers declining registration and collaboration with the Penn Telegenetics Program (n=56)

| Reason for Provider Registration Refusal | Number of occurrences*<br>N (%) |
| --- | --- |
| Provider reported they would facilitate genetic services locally. In some cases they spoke with the patient directly about the options and they both agreed to a preference for local services. | 16 (22.5) |
| Provider reported they were not comfortable being on the test order with the Penn Telegenetics GC. | 16 (22.5) |
| Patient changed their mind after discussing genetic testing with the provider. | 10 (14.1) |
| Office reported that the provider would not register until the patient was seen (most had not been seen recently) or until they could have a discussion with the patient. In some cases they were willing to do this by phone but in most they required the patient to come in for an in-person visit. | 10 (14.1) |
| Despite multiple attempts the research team and physician lead could not reach the provider to discuss their reason for refusing registration and collaboration. | 7 (12.5) |
| Provider no longer was affiliated with practice (e.g. retired or left the practice). Patients then needed to identify a new provider or see a new provider in the practice. | 6 (10.7) |
| Unknown (patient or provider could not be reached) | 5 (8.9) |
| Provider delayed (no outright refusal) and by the time they registered the patient was no longer interested or could not be reached. | 1 (1.8) |

\*Providers could be coded for more than one reason for refusal

Among registered providers, we did not receive any requests from providers for additional information about test results or recommendations. We also did not receive subsequent referrals for testing in new patients in provider practices.

These data demonstrate that the collaborative in-home PCP/LP telegenetics delivery model is feasible and has potential to improve access to genetic testing across the US. Our data demonstrate that the large majority of local providers are willing to register with the program and provide access to genetic testing for their patient. The success might be due to some PCPs interest in testing for their patients but low comfort or training in genetics.^10–12^ Willingness may also have been related to the low burden for the local practice.^11^ Future work to understand local providers motivations, perceptions and experience engaging with telegenetics programs could be helpful.

Still, we did encounter a subset of PCPs that declined to collaborate with the telegenetics program. Some providers refused given a preference for using local genetic testing services, although we don’t know if patients ultimately completed testing. Some reported not being comfortable being on the test requisition. It may be useful to develop materials for the patient to share with the PCP/LPs when discussing the testing process. This may prompt mutual informed decision making between patient and provider, greater willingness for providers to collaborate and subsequent testing. Interviews with local providers could provide additional insights and areas for improvement.

Representing a potential missed opportunity, we have not received follow-up contact from local providers, nor have registered providers referred other patients. We had hoped that providers would see the service as a low-burden for them and of benefit for their individual patients. It could be that providers felt comfortable modifying medical care based on the genetic test results. or it may not be impacting care at all. Unfortunately, we don’t know at this time how providers used the results or GC notes that we sent.^13^ Several studies are collecting longitudinal behaviors, which could provide insights when these data are available. Limited referrals and engagement could be due to time constraints in busy local practices^14,15^, limited interest in genetic testing^16^, or lack of training in genetic medicine^17^ or identification of ideal candidates for genetic testing^11,12,18^.

While the program has been successful, we believe there are areas for improvement. Future qualitative research will include interviews with providers to understand their perceptions of the program, and their perceived barriers and facilitators to testing uptake. It will also be important to understand their perceived utility of genetic testing, and how it impacts their local workflow. Interviews may also provide insights on how we can improve our collaborative model to better address provider questions and reservations. Connecting with other practice members (e.g. nurses or office staff) or providing educational content for providers could be beneficial to increasing uptake of the program.

These data demonstrate that most local providers are willing to collaborate with a centralized cancer genetics program to provide remote genetic services to their patients in the home.

Refining procedures or increasing engagement may provide opportunities to increase willingness among local providers and to facilitate additional referrals and access to genetic services in community practices.

## Data Availability

All data produced in the present work are contained in the manuscript

## Notes

### Competing Interest Statement

The authors have declared no competing interest.

### Funding Statement

Research reported in this publication was supported by the National Human Genome Research Institute of the National Institutes of Health under Award Number K00HG012487. The content is solely the responsibility of the authors and does not necessarily represent the official views of the National Institutes of Health

### Author Declarations

The Institutional Review Board of the University of Pennsylvania gave ethical approval for this work.

